# Cumulative exposure to risk factors and subsequent cardiovascular disease events: The Jackson Heart Study

**DOI:** 10.64898/2026.07.27.26359076

**Authors:** Rishi V. Parikh, Paul Muntner, Shakia T. Hardy, Mario Sims, Yuan-I Min, Kendra D. Sims, Alexis Reeves, Yongmei Li, Michelle C. Odden

## Abstract

**Background:** Cumulative exposure to risk factor levels may capture cardiovascular disease (CVD) risk independent of a single measurement. We examined the association between cumulative exposure to systolic blood pressure (SBP), low-density lipoprotein cholesterol (LDL-C), and fasting plasma glucose (FPG) with CVD events among African American participants in the Jackson Heart Study (JHS).

**Methods:** We included all JHS participants with SBP, LDL-C, and FPG measurements at Exams 1 (2000-2004) and 3 (2009-2013) who were alive at 8 years of follow-up after Exam 1. We calculated 8-year cumulative exposures as the area under the trajectory of each risk factor using all available measurements from Exams 1, 2, and 3. Adjudicated coronary heart disease (CHD), stroke, and heart failure (HF) events after the 8-year exposure period and through December 31, 2016 were evaluated separately using conditional Cox regression models with each cumulative exposure, respective values at 8 years, and Exam 1 covariates as independent variables. Models were conducted separately for each risk factor, and exposures were centered and scaled for comparability.

**Results:** Among 3188 eligible JHS participants, mean age was 54 years, and 64% were women. Higher cumulative exposures to SBP and LDL-C were associated with CHD events after adjustment for covariates and the 8-year values [adjusted hazard ratios (aHR) per SD: 1.37 per 136 mmHg-years of SBP (95% CI: 1.09, 1.71), and 1.56 per 309 mg/dL-years of LDL-C (95% CI: 1.02, 2.37)]. Higher cumulative exposure to SBP and FPG were associated with HF events after adjustment [aHR: 1.15 per 136 mmHg-years of SBP (95% CI: 1.04, 1.27) and 1.10 per 262 mg/dL-years of FPG (95% CI: 1.02, 1.18)]. Only the 8-year values, but not cumulative exposures, were associated with stroke risk after full adjustment.

**Conclusions:** Cumulative exposure to risk factors may provide additional information on CVD risk compared with single measurements later in life, particularly for CHD and HF events.

**Clinical Perspective:** *What is new?:*

- We examined associations between cardiovascular risk factors (systolic blood pressure [SBP], low density lipoprotein cholesterol [LDL-C], fasting plasma glucose [FPG]) and coronary heart disease (CHD), heart failure (HF), and stroke in a population of African American adults.
- We calculated cumulative exposures as the area under the trajectory of each risk factor over 8 years and compared associations with a single value measured at the end of the exposure period.
- Cumulative exposure to SBP, LDL-C and FPG conferred additional risk for CHD and HF events over single values.

*What are the clinical implications?:*

- Long-term exposure to both clinical and subclinical levels of risk factors may improve comprehensive CVD risk assessment in African-American adults.
- These findings emphasize the importance of regular monitoring and sustained low levels of SBP, LDL-C, and FPG to manage lifetime CVD risk.
- Interventions aimed at lowering and managing exposure to risk factors over the life course rather than solely treating risk factors once they are high, may represent effective strategies for reducing the burden of CVD in African American adults.

## Introduction

Hypertension, dyslipidemia, and diabetes are three of the most important risk factors for the development and progression of cardiovascular disease (CVD), and all have been shown to be modifiable with medication and lifestyle modification.^1^ Compared to non-Hispanic White adults, African American adults have a higher prevalence of hypertension and diabetes, as well as a higher incidence of heart failure (HF) and stroke ^2^ These disparities are more prominent at younger ages, leading to a higher cumulative lifetime burden of risk factors among African American individuals.^2^ Moreover, long-term exposure to high blood pressure and cholesterol has been associated with the progression of atherosclerosis beyond a single measurement.^3,4^ Therefore, systolic blood pressure (SBP), cholesterol, or glucose measured at a single point may not fully capture CVD risk, especially in African American adults.

Long-term and early exposure to high SBP is well known to confer an increased risk of coronary artery calcification^3^, cardiac dysfunction^5^, CVD events, and mortality^6^ in both African American and non-Hispanic White adults. However, the cumulative effect of exposure to high low-density lipoprotein cholesterol (LDL-C) in African American adults is less well-documented.^7^ Paradoxically, African American adults have a lower population prevalence, but a higher age-specific incidence, of hypercholesterolemia compared with non-Hispanic White adults; this may suggest that LDL-C plays a stronger role in CVD development and related mortality in African Americans, leading to survivor bias in the lower prevalence estimates.^2,8^ Furthermore, although long-term exposure to high blood glucose levels is associated with increased risk of diabetes^9^ and microvascular complications,^10^ few studies have examined the impact of cumulative exposure to high blood glucose levels on CVD risk in any population.^11,12^

Given the high prevalence of diabetes and hypertension in the African American adult population, and the relatively lower prevalence of hypercholesterolemia, evaluating the relative importance and synergistic effect of risk factors for CVD in African American individuals can inform prevention strategies. In addition, focusing on the effects of cumulative and longitudinal exposure to CVD risk factors can help inform optimal health maintenance activities across the lifespan through more accurate assessment of long-term risk. In this study, we analyzed data from participants of the Jackson Heart Study (JHS) to evaluate the associations between 8-year cumulative exposures to SBP, cholesterol, and blood glucose and subsequent CHS, stroke, or HF events. We hypothesized that cumulative exposure to each risk factor will be more strongly associated with each type of CVD event compared to more recent measurements and other risk factors.

## Methods

The data used in this study can be made available to other researchers for purposes of reproducing the results or replicating the procedure by following the JHS publications procedures and data use agreements.

### Study Participants

The JHS is a community-based cohort study evaluating risk factors for cardiovascular and related diseases among African American adults residing in the Jackson, Mississippi metropolitan area. Details of the JHS study design, recruitment, and data collection have been described previously.^13^ Briefly, data and biologic materials were collected from 5,306 participants who provided medical and psychosocial histories and had an array of physical and biochemical measurements and diagnostic procedures during a baseline examination (2000-2004) and two follow-up examinations (2005-2008 and 2009-2013). Annual follow-up interviews and cohort surveillance of cardiovascular events and mortality are continuing, and a fourth examination is in progress. The JHS was approved by the institutional review boards of Jackson State University, Tougaloo College, and the University of Mississippi Medical Center in Jackson, Mississippi. All study participants provided written informed consent.

For this study, we excluded JHS participants who did not attend Exam 3 (n=1487), had missing values for SBP, LDL-C, and fasting plasma glucose (FPG) at Exams 1 or 3 (n=625), or who had died within 8 years after Exam 1 (n=6).

### Risk factors and cumulative exposures

At Exam 1, two seated SBP and diastolic blood pressures were measured in the right arm of participants using a random-zero sphygmomanometer (RZS, Hawksley and Sons Limited). At Exams 2 and 3, blood pressures were measured using an oscillometric device (OD, Omron HEM-907XL). Blood pressures measured using the RZS were standardized to the OD.^14^ The first blood pressure was obtained after allowing the participant to rest for 5 minutes in a seated position, and the second blood pressure was obtained after waiting 1 additional minute. The average of the 2 measurements was used. We focus on SBP rather than diastolic blood pressure in this study, as it has a stronger association with CVD risk. Fasting serum LDL-C and plasma glucose were assayed using standard techniques and LDL-C was calculated by the Friedewald equation.^15^ In a secondary analysis, we also analyzed non-high-density lipoprotein (non-HDL) cholesterol as the difference between total and HDL cholesterol. Plasma total and HDL cholesterol concentrations were measured using standard enzymatic methods, on a Vitros 950 or 250, Ortho-Clinical Diagnostics analyzer (Raritan, NJ) in accordance with the College of American Pathologists Proficiency Testing Program.

Cumulative exposures were calculated as the area under the curve (AUC), or the trajectory, for each individual in the first 8 years after Exam 1 using data from measurements at Exam 1, 2 (if available), and 3 (**Figure 1A**). The 8-year value for each measurement was either interpolated (i.e., if Exam 3 occurred after the 8-year index date) or extrapolated (i.e., if Exam 3 occurred before the 8-year index date) using the trajectory from the previous exam to the Exam 3 value. The 8-year index date was chosen as it was the median time from Exam 1 to Exam 3 in the JHS cohort, so approximately equal numbers of participants had 8-year values interpolated (n=1556) and extrapolated (n=1632). In sensitivity analyses, we also considered two formulations of cumulative exposure to levels above clinically relevant thresholds (i.e., SBP ≥ 130 mmHg, LDL-C ≥ 130 mg/dL, FPG ≥ 126 mg/dL): 1) the AUC above the threshold value only, and 2) the proportion of time spent above the threshold value.

**Figure 1.**
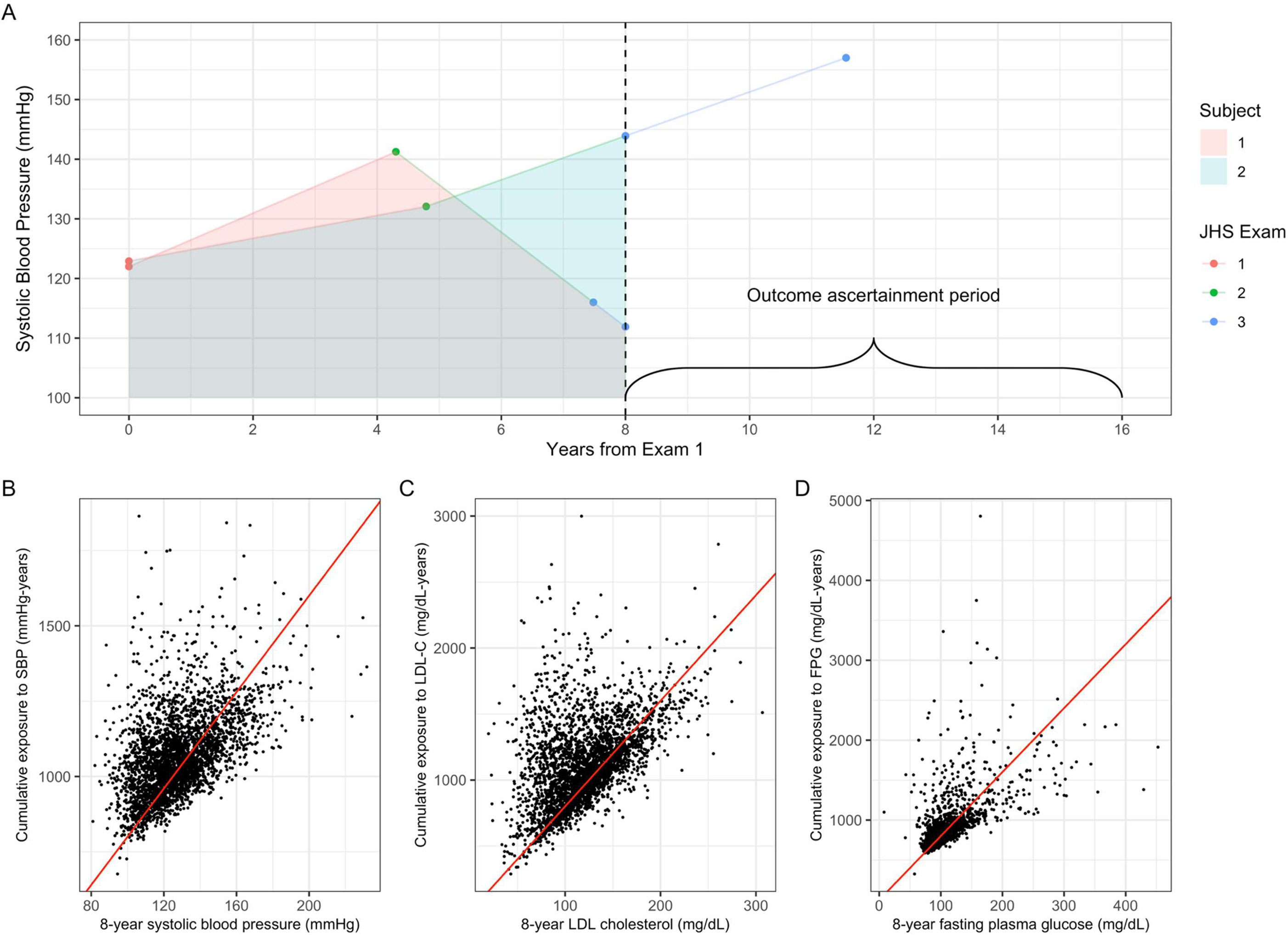
Cumulative exposures to risk factors compared to single time point measurements in the Jackson Heart Study. **A**: Sample cumulative exposure calculation for 2 subjects. Cumulative exposures were calculated across the first 8 years after Exam 1 as the area under the trajectory using the trapezoid rule. The 8-year value was interpolated (Subject 2) or extrapolated (Subject 1) based on the timing of the Exam 3 measurement (assigned denoted by the blue dots intersecting with the vertical dotted line). CVD events were assessed from the 8-year index date through December 31, 2016 (max of 8.3 years of follow-up). **B, C, D**: Cumulative exposures vs. 8-year values for systolic blood pressure (B), low-density lipoprotein cholesterol (C), and fasting plasma glucose (C). The red line represents the cumulative exposure assuming constant exposure.

### Outcomes

CVD events included fatal or non-fatal coronary heart disease (CHD), HF and stroke events (including recurrent events) starting from 8 years after the Exam 1 date through December 31, 2016. Methods for defining CVD events in the JHS evolved from the Atherosclerosis Risk in Communities (ARIC) study and have been previously described.^16,17^ Trained and certified personnel performed surveillance and monitoring of CHD, stroke, and HF. After participants were contacted by telephone to identify health events including diagnostic tests, hospitalizations, or death, the medical record abstraction unit obtained data including discharge lists and death certificates from hospitals and state offices for verification. Eligible events were classified as definite or probable fatal or nonfatal CHD, HF, and stroke events including recurrent events by a computer algorithm and follow-up review and adjudication by 2 independent physician reviewers.

### Covariates

All covariates were measured at Exam 1. Height and waist circumference were measured by study personnel, and waist-to-height ratio was calculated by dividing waist circumference by weight. Serum creatinine was measured using the rate Jaffe reaction, and the kidney function was assessed using the estimated glomerular filtration rate (eGFR) calculated using the 2009 Chronic Kidney Disease Epidemiology Collaboration (CKD-EPI) equation.^18^ Information on diabetes, hypertension, CVD history, medication use, alcohol use within the past 12 months, current smoking status (yes/no), income level including poor, lower-middle, upper-middle, high, based on income adjusted for family size and year, and educational attainment including less than high school, high school/GED, additional schooling self-reported using standardized questionnaires during the baseline examination. Missing data for income (n=459) was treated as a separate category, and missing data for educational attainment (n=4) were imputed using the mode value. Missing data for waist-to-height ratio (n=2) and eGFR (n=1) were imputed using the mean values.

### Statistical Analysis

All analyses were conducted in R Version 4.5.1 (R Core Team, 2025). We first described characteristics of the analytic cohort at Exam 1 and compared these characteristics to those excluded from our analyses in secondary analyses. We calculated crude rates per 100 person-years and 95% Poisson confidence intervals (CI) for each outcome from the 8-year index date through the end of follow-up by quartile of each cumulative exposure measure. To evaluate associations between each cumulative exposure measurement and each outcome, we used Cox regression models conditional on each recurrent event.^19^ We adjusted for the following confounders measured at Exam 1 and identified *a priori*: age, sex, income level, educational attainment, current smoking, alcohol use, waist-to-height ratio, total cholesterol, triglycerides, eGFR, prior CVD, diabetes, antihypertensive medication use, glucose-lowering medication use, and statin use. Also, we included the corresponding 8-year value for each exposure and reported the hazard ratio (HR) and 95% CI associated with both the cumulative exposure and the 8-year value. All variables were standardized by subtracting the mean and dividing by the standard deviation; therefore, all hazard ratios can be interpreted as the change in relative hazard of the outcome per increase in 1 standard deviation. Finally, we conducted a combined model for each outcome, which included all three cumulative exposures (SBP, LDL-C, and FPG) and 8-year values along with covariates. To examine the association between measures and incident events only, we performed sensitivity analyses excluding those with CVD prior to the 8-year index date and analyzing the time to first event instead of all recurrent events.

## Results

### Cohort characteristics

Among 5306 participants in the JHS, 3188 attended and had risk factor measurements at both Exams 1 and 3; of these, 2988 participants had measurements at Exam 2. Mean age at Exam 1 was 53.6 years, 64% were women, and 10.9% were smokers (**Table 1**). Mean (SD) SBP was 126 (15.8) mm Hg; 51.5% of participants reported a diagnosis of hypertension, and 47.5% were taking antihypertensive medication. Mean (SD) LDL-C was 127 (36.1) mg/dL, and 11.5% were taking a statin. Mean (SD) FPG was 96.1 (24.7) mg/dL, with 15% reporting a diagnosis of diabetes and 8.1% taking medication for diabetes. In addition, 7.2% of participants reported a history of CVD prior to Exam 1. Characteristics of participants by quartiles of LDL-C and FPG are presented in **Tables S1-S2**. Excluded participants were less likely to report currently using alcohol, ever smoking, be of lower socioeconomic status, and more likely to have CVD risk factors such as hypertension and diabetes (**Table S3**).

**Table 1.** Characteristics of JHS participants at Exam 1, overall and stratified quartile of cumulative systolic blood pressure.

| Exam 1 variables | Overall<br>(N=3188) | Quartiles of cumulative systolic blood pressure |  |  |  |
| --- | --- | --- | --- | --- | --- |
|  |  | Q1: 677-<976<br>mmHg-years<br>(N=797) | Q2: 976-<1050<br>mmHg-years<br>(N=797) | Q3: 1050-<1137<br>mmHg-years<br>(N=797) | Q4: 1138-1864<br>mmHg-years<br>(N=797) |
| Demographics |  |  |  |  |  |
| Age, years | 53.6 (12.0) | 47.0 (11.7) | 52.5 (11.1) | 55.4 (11.0) | 59.3 (10.9) |
| Female sex, n (%) | 2047 (64.2%) | 544 (68.3%) | 484 (60.7%) | 511 (64.1%) | 508 (63.7%) |
| Alcohol use, n (%) | 1539 (48.3%) | 429 (53.8%) | 400 (50.2%) | 376 (47.2%) | 334 (41.9%) |
| Current smoker, n (%) | 349 (10.9%) | 81 (10.2%) | 96 (12.0%) | 68 (8.5%) | 104 (13.0%) |
| Ever smoker, n (%) | 920 (28.9%) | 205 (25.7%) | 234 (29.4%) | 203 (25.5%) | 278 (34.9%) |
| Anthropometrics |  |  |  |  |  |
| Weight, kgs | 90.6 (21.3) | 89.4 (21.1) | 90.0 (19.6) | 91.9 (22.0) | 91.1 (22.3) |
| Missing, n (%) | 2 (0.1%) | 0 (0%) | 1 (0.1%) | 0 (0%) | 1 (0.1%) |
| Height, cm | 169 (9.26) | 169 (8.91) | 170 (9.33) | 169 (9.25) | 169 (9.50) |
| Missing, n (%) | 2 (0.1%) | 0 (0%) | 1 (0.1%) | 0 (0%) | 1 (0.1%) |
| Waist circumference, cm | 100 (16.1) | 97.2 (15.8) | 99.2 (15.0) | 101 (16.5) | 102 (16.5) |
| Missing, n (%) | 3 (0.1%) | 0 (0%) | 1 (0.1%) | 0 (0%) | 2 (0.3%) |
| Waist-to-height ratio | 0.593 (0.0969) | 0.575 (0.0948) | 0.586 (0.0906) | 0.603 (0.0997) | 0.606 (0.0990) |
| Missing, n (%) | 3 (0.1%) | 0 (0%) | 1 (0.1%) | 0 (0%) | 2 (0.3%) |
| Medical history, n (%) |  |  |  |  |  |
| Antihypertensive use | 1514 (47.5%) | 224 (28.1%) | 357 (44.8%) | 433 (54.3%) | 500 (62.7%) |
| Statin use | 361 (11.3%) | 66 (8.3%) | 91 (11.4%) | 94 (11.8%) | 110 (13.8%) |
| Beta blocker use | 278 (8.7%) | 42 (5.3%) | 44 (5.5%) | 84 (10.5%) | 108 (13.6%) |
| Calcium channel blocker use | 544 (17.1%) | 65 (8.2%) | 106 (13.3%) | 166 (20.8%) | 207 (26.0%) |
| Diuretic use | 934 (29.3%) | 147 (18.4%) | 235 (29.5%) | 261 (32.7%) | 291 (36.5%) |
| Glucose-lowering therapy | 259 (8.1%) | 39 (4.9%) | 56 (7.0%) | 73 (9.2%) | 91 (11.4%) |
| Hypertension | 1641 (51.5%) | 197 (24.7%) | 334 (41.9%) | 435 (54.6%) | 675 (84.7%) |
| Diabetes | 489 (15.3%) | 85 (10.7%) | 107 (13.4%) | 139 (17.4%) | 158 (19.8%) |
| Cardiovascular disease | 35 (4.4%) | 38 (4.8%) | 61 (7.7%) | 95 (11.9%) | 229 (7.2%) |
| **Vitals and laboratory values** |  |  |  |  |  |
| Systolic blood pressure, mmHg | 126 (15.8) | 110 (6.94) | 121 (6.81) | 129 (8.18) | 144 (15.0) |
| LDL cholesterol, mg/dL | 127 (36.1) | 124 (34.6) | 126 (36.3) | 127 (36.6) | 133 (36.5) |
| Fasting plasma glucose, mg/dL | 96.1 (24.7) | 91.9 (20.1) | 95.3 (23.7) | 96.9 (26.7) | 100 (27.0) |
| HDL cholesterol, mg/dL | 52.0 (14.3) | 51.4 (13.4) | 51.0 (13.5) | 52.9 (14.3) | 52.9 (15.7) |
| Total cholesterol, mg/dL | 199 (38.9) | 193 (36.9) | 197 (38.5) | 200 (38.8) | 207 (40.0) |
| Triglycerides, mg/dL | 99.5 (52.1) | 89.7 (47.6) | 98.2 (54.0) | 102 (53.0) | 108 (51.8) |
| Estimated glomerular filtration rate,<br>mL/min/1.73m<sup>2</sup> | 96.5 (19.7) | 103 (19.9) | 97.0 (19.0) | 95.0 (18.7) | 91.3 (19.3) |
| **Social risk factors** |  |  |  |  |  |
| Health insurance type, n (%) |  |  |  |  |  |
| Uninsured | 398 (12.5%) | 95 (11.9%) | 111 (13.9%) | 92 (11.5%) | 100 (12.5%) |
| Private only | 1995 (62.6%) | 588 (73.8%) | 530 (66.5%) | 497 (62.4%) | 380 (47.7%) |
| Public only | 416 (13.0%) | 63 (7.9%) | 77 (9.7%) | 101 (12.7%) | 175 (22.0%) |
| Private and Public | 368 (11.5%) | 48 (6.0%) | 76 (9.5%) | 104 (13.0%) | 140 (17.6%) |
| Missing, n (%) | 11 (0.3%) | 3 (0.4%) | 3 (0.4%) | 3 (0.4%) | 2 (0.3%) |
| Income category, n (%) |  |  |  |  |  |
| Poor | 304 (9.5%) | 81 (10.2%) | 64 (8.0%) | 68 (8.5%) | 91 (11.4%) |
| Lower-middle | 612 (19.2%) | 126 (15.8%) | 142 (17.8%) | 161 (20.2%) | 183 (23.0%) |
| Upper-middle | 853 (26.8%) | 223 (28.0%) | 209 (26.2%) | 233 (29.2%) | 188 (23.6%) |
| Affluent | 960 (30.1%) | 241 (30.2%) | 272 (34.1%) | 244 (30.6%) | 203 (25.5%) |
| Missing, n (%) | 459 (14.4%) | 126 (15.8%) | 110 (13.8%) | 91 (11.4%) | 132 (16.6%) |
| Educational attainment, n (%) |  |  |  |  |  |
| Less than high school | 434 (13.6%) | 72 (9.0%) | 85 (10.7%) | 107 (13.4%) | 170 (21.3%) |
| High school graduate/GED | 607 (19.0%) | 123 (15.4%) | 157 (19.7%) | 151 (18.9%) | 176 (22.1%) |
| Attended vocational school, trade school,<br>or college | 2143 (67.2%) | 599 (75.2%) | 555 (69.6%) | 539 (67.6%) | 450 (56.5%) |
| Missing, n (%) | 4 (0.1%) | 3 (0.4%) | 0 (0%) | 0 (0%) | 1 (0.1%) |
| **Physical activity** |  |  |  |  |  |
| Sport index | 2.29 (1.31) | 2.47 (1.33) | 2.33 (1.32) | 2.26 (1.27) | 2.09 (1.29) |
| Missing, n (%) | 55 (1.7%) | 10 (1.3%) | 13 (1.6%) | 16 (2.0%) | 16 (2.0%) |
| Home/Yard index | 2.32 (0.614) | 2.34 (0.618) | 2.32 (0.619) | 2.30 (0.598) | 2.32 (0.623) |
| Missing, n (%) | 30 (0.9%) | 5 (0.6%) | 4 (0.5%) | 7 (0.9%) | 14 (1.8%) |
| Active living index | 2.14 (0.798) | 2.23 (0.780) | 2.17 (0.811) | 2.11 (0.793) | 2.04 (0.798) |
| Missing, n (%) | 10 (0.3%) | 4 (0.5%) | 1 (0.1%) | 2 (0.3%) | 3 (0.4%) |

Participants had a median 8-year cumulative exposure to SBP of 1050 (IQR: 976, 1138) mm Hg-years; this is equivalent to an average SBP of 131 mm Hg. Median cumulative exposure to LDL-C was 1082 (IQR: 899, 1281) mg/dL-years which is equivalent to an average LDL-C of 135 mg/dL, and median cumulative exposure to FPG was 775 (IQR: 724, 855) mg/dL-years which is equivalent to an averageFPG of 97 mg/dL. The median 8-year SBP was 125 (IQR: 114, 138) mm Hg, LDL-C was 118 (IQR: 95, 143) mg/dL, and FPG was 97 (89, 104). Pearson correlation coefficients for cumulative exposure measures with 8-year values were 0.49 for SBP, 0.47 for LDL-C, and 0.63 for FPG **(Figures 1B-1D).**

### Primary outcome analyses

After the 8-year index date, participants had a median (25th, 75^th^ percentile) follow-up of 6.2 (5.3, 6.8) years. There were a total of 88 CHD, 519 HF, and 78 stroke events corresponding to 74, 189, and 70 individuals, respectively. Crude incidence rates of CHD, HF, and stroke events were higher across increasing quartiles of all cumulative exposures (**Table 2, Table S4**).

**Table 2.**
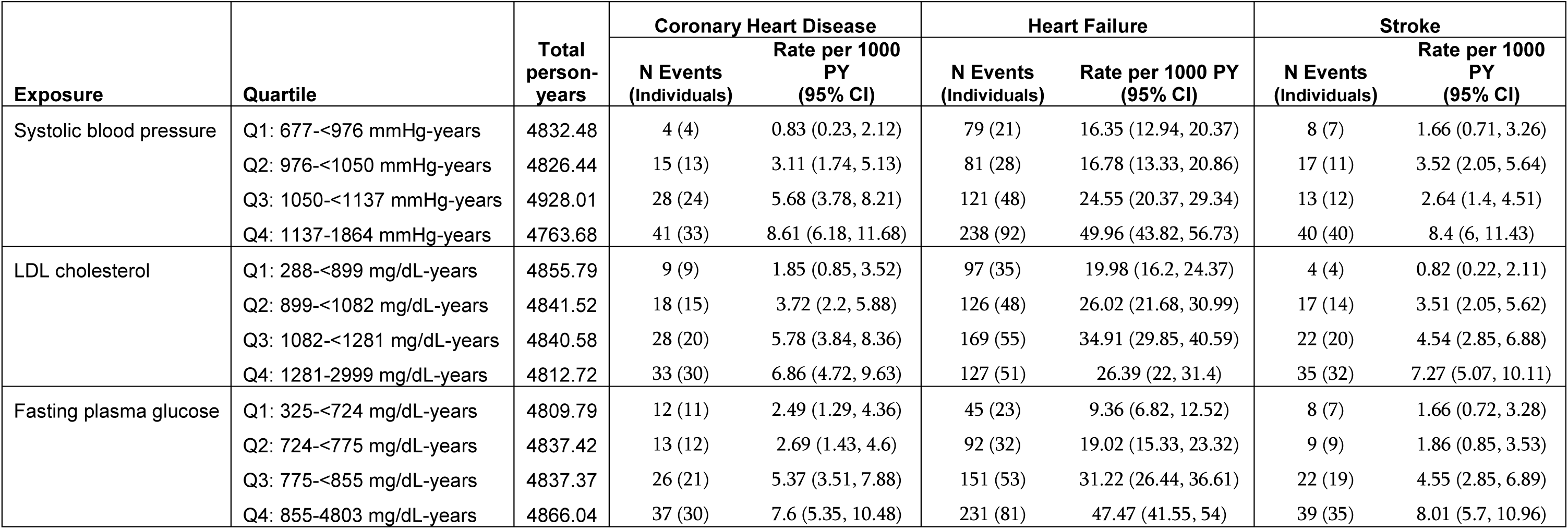
Crude rates (95% CI) per 100 person-years of cardiovascular events after Exam 3, by quartile of each cumulative exposure measurement.

| Exposure | Quartile | Total person-years | Coronary Heart Disease |  | Heart Failure |  | Stroke |  |
| --- | --- | --- | --- | --- | --- | --- | --- | --- |
|  |  |  | N Events (Individuals) | Rate per 1000 PY (95% CI) | N Events (Individuals) | Rate per 1000 PY (95% CI) | N Events (Individuals) | Rate per 1000 PY (95% CI) |
| Systolic blood pressure | Q1: 677-<976 mmHg-years | 4832.48 | 4 (4) | 0.83 (0.23, 2.12) | 79 (21) | 16.35 (12.94, 20.37) | 8 (7) | 1.66 (0.71, 3.26) |
|  | Q2: 976-<1050 mmHg-years | 4826.44 | 15 (13) | 3.11 (1.74, 5.13) | 81 (28) | 16.78 (13.33, 20.86) | 17 (11) | 3.52 (2.05, 5.64) |
|  | Q3: 1050-<1137 mmHg-years | 4928.01 | 28 (24) | 5.68 (3.78, 8.21) | 121 (48) | 24.55 (20.37, 29.34) | 13 (12) | 2.64 (1.4, 4.51) |
|  | Q4: 1137-1864 mmHg-years | 4763.68 | 41 (33) | 8.61 (6.18, 11.68) | 238 (92) | 49.96 (43.82, 56.73) | 40 (40) | 8.4 (6, 11.43) |
| LDL cholesterol | Q1: 288-<899 mg/dL-years | 4855.79 | 9 (9) | 1.85 (0.85, 3.52) | 97 (35) | 19.98 (16.2, 24.37) | 4 (4) | 0.82 (0.22, 2.11) |
|  | Q2: 899-<1082 mg/dL-years | 4841.52 | 18 (15) | 3.72 (2.2, 5.88) | 126 (48) | 26.02 (21.68, 30.99) | 17 (14) | 3.51 (2.05, 5.62) |
|  | Q3: 1082-<1281 mg/dL-years | 4840.58 | 28 (20) | 5.78 (3.84, 8.36) | 169 (55) | 34.91 (29.85, 40.59) | 22 (20) | 4.54 (2.85, 6.88) |
|  | Q4: 1281-2999 mg/dL-years | 4812.72 | 33 (30) | 6.86 (4.72, 9.63) | 127 (51) | 26.39 (22, 31.4) | 35 (32) | 7.27 (5.07, 10.11) |
| Fasting plasma glucose | Q1: 325-<724 mg/dL-years | 4809.79 | 12 (11) | 2.49 (1.29, 4.36) | 45 (23) | 9.36 (6.82, 12.52) | 8 (7) | 1.66 (0.72, 3.28) |
|  | Q2: 724-<775 mg/dL-years | 4837.42 | 13 (12) | 2.69 (1.43, 4.6) | 92 (32) | 19.02 (15.33, 23.32) | 9 (9) | 1.86 (0.85, 3.53) |
|  | Q3: 775-<855 mg/dL-years | 4837.37 | 26 (21) | 5.37 (3.51, 7.88) | 151 (53) | 31.22 (26.44, 36.61) | 22 (19) | 4.55 (2.85, 6.89) |
|  | Q4: 855-4803 mg/dL-years | 4866.04 | 37 (30) | 7.6 (5.35, 10.48) | 231 (81) | 47.47 (41.55, 54) | 39 (35) | 8.01 (5.7, 10.96) |

Cumulative exposure to SBP was more strongly associated with CHD (HR per 1 SD [136 mmHg-years]: 1.37, 95% CI: 1.09, 1.71) and HF (HR: 1.15, 95% CI: 1.04, 1.27) than the 8-year values [HR per 1 SD (18.8 mmHg) for CHD: 1.03, 95% CI: 0.84, 1.27; and HF: 0.96, 95% CI: 0.87, 1.06]; corresponding associations with stroke were not statistically significant (HR: 1.22, 95% CI: 0.95, 1.57), (**Figure 2, Table S5**).

**Figure 2.**
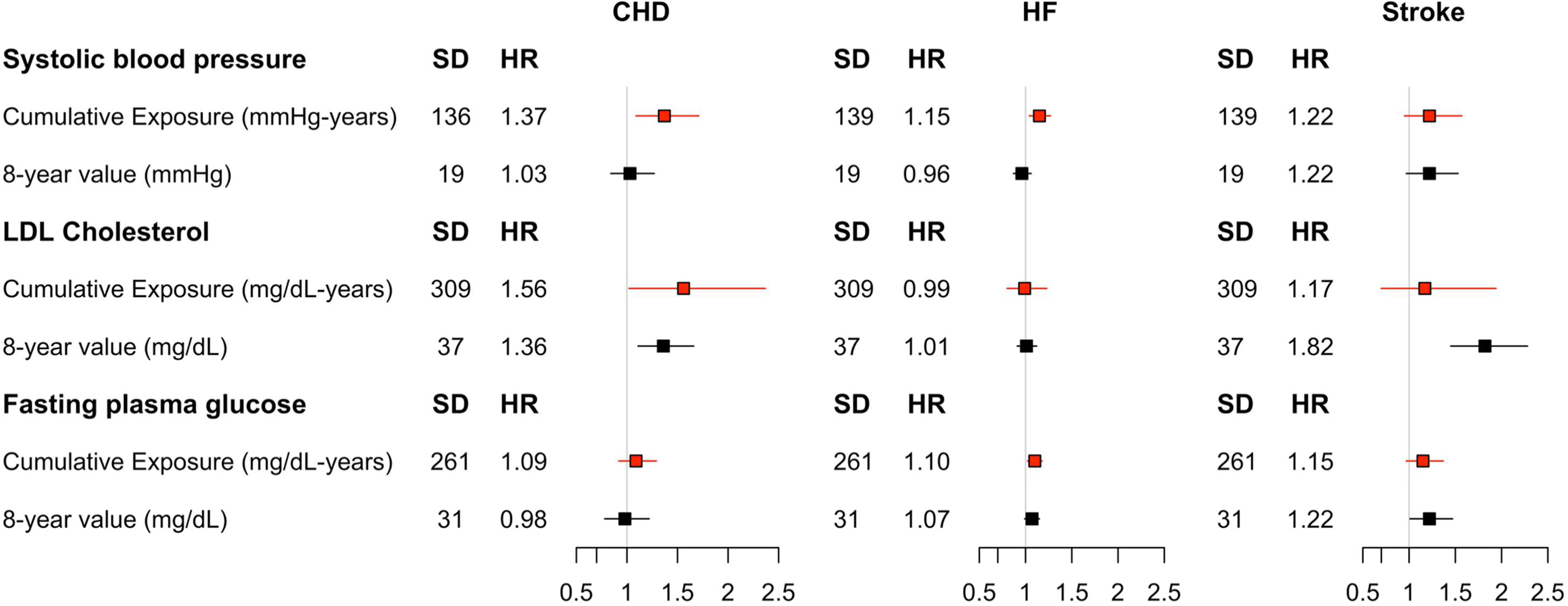
Hazard ratios for the association between cumulative exposures of risk factors and CVD events in the Jackson Heart Study. Covariates include: Age, sex, income, education, current smoker, alcohol use, waist-to-height ratio, total cholesterol, triglycerides, eGFR, CVD history, diabetes, anti-hypertensive medication use, glucose-lowering therapy, and statin use.

Cumulative exposure to LDL-C was associated with CHD [HR per 1 SD (309 mg/dL-years): 1.56, 95% CI: 1.02, 2.37] along with the 8-year LDL-C value (HR: 1.36, 95% CI: 1.11, 1.66). Neither the cumulative exposure nor the 8-year LDL-C were associated with HF, but the 8-year LDL-C value was associated with stroke (HR: 1.82, 95% CI: 1.45, 2.28) while the cumulative exposure was not (HR: 1.17, 95% CI: 0.70, 1.94) (**Figure 2, Table S5**). Results for non-HDL-C are shown in **Table S5**.

Higher cumulative exposure to FPG and the 8-year value were both associated with higher risk for all outcomes (**Figure 2, Table S5**). Higher cumulative FPG was associated with HF [cumulative HR per 1 SD (262 mg/dL-years): 1.10, 95% CI: 1.02, 1.18; 8-year value HR: 1.07, 95% CI: 0.99, 1.15], the 8-year value was associated with stroke (cumulative HR: 1.15, 95% CI: 0.97, 1.37; 8-year HR: 1.22, 95% CI: 1.01, 1.47), and neither measure was associated with CHD (cumulative HR: 1.09, 95% CI: 0.92, 1.29; 8-year HR: 0.98, 95% CI: 0.78, 1.22).

### Combined model and sensitivity analyses

For all exposures, the calculation of cumulative exposure using the total AUC (i.e., above 0) displayed point estimates that were not statistically different compared to the AUC above a threshold or the proportion of time spent above a threshold (**Table S5**).

In a fully adjusted, combined model for CHD including cumulative exposures to SBP, LDL-C, FPG, the respective 8-year values, and covariates, higher cumulative exposure to SBP (HR: 1.35, 95% CI: 1.11, 1.65) and LDL-C (HR: 1.64, 95% CI: 1.08, 2.47) along with the 8-year LDL-C (HR: 1.30, 95% CI: 1.06, 1.58) were associated with CHD events (**Table S6**). In the combined model for HF, higher cumulative exposure to SBP (HR: 1.10, 95% CI: 1.01, 1.20) and FPG (HR: 1.09, 95% CI: 1.01, 1.17) were associated with HF. In the combined model for stroke, higher cumulative exposure to FPG (HR: 1.24, 95% CI: 1.05, 1.47) and the 8-year LDL-C (HR: 1.67, 95% CI: 1.34, 2.08) were associated with stroke.

In a sensitivity analysis of time to first event among those with no CVD at Exam 1, there were 2959 participants and 57 CHD, 148 HF, and 59 stroke events. In both single models for each exposure (**Table S7**) and combined models with all exposures (**Table S8**), cumulative exposure to SBP and LDL-C and the 8-year LDL-C were associated with CHD, cumulative exposure to FPG was associated with HF, and the 8-year LDL-C was associated with stroke.

## Discussion

In this community-based cohort of African American adults, cumulative SBP and LDL-C were associated with higher rates of CHD, and cumulative SBP and FPG were modestly associated with HF. In contrast, a single value of LDL-C was more strongly associated with stroke than the cumulative exposure. These results suggest that cumulative exposure measurements of some cardiovascular risk factors may provide additional information on CVD event risk compared to single measures in African American adults, especially for CHD and HF events. Using single measurements alone may underestimate this risk.

The current findings align with prior reports of a strong association between cumulative SBP exposure and multiple CVD endpoints, including CHD, HF, and stroke,^5,15,16^ and confirm these results in a large population of African American adults. We also confirm prior findings from Zhang et al.,^7^ who observed a strong association between cumulative LDL-C and CHD, but not HF or stroke. In their study, which pooled data from four prospective cohorts including ∼18,000 participants, the authors reported that point estimates for the association between cumulative LDL-C and CHD were lower in African American than non-Hispanic White participants, although they were unable to examine this in more detail due to the low representation of African American participants. In contrast, in a larger population of exclusively African American adults, both higher cumulative exposure to LDL-C and a higher later-life LDL-C value were strongly associated with CHD risk in the JHS population, after adjustment for SBP and other risk factors. These findings highlight the importance of LDL-C as a risk factor for CHD and the need for early and ongoing lipid management in African American adults, despite other epidemiologic evidence suggesting relatively lower prevalences of hypercholesterolemia at a population level and studies of earlier follow-up periods in JHS (2000 – 2008) showing weaker associations.^22,23^ LDL-C was the risk factor most strongly associated with stroke events; however, contrary to our hypothesis, the proximal 8-year value was more strongly associated than the cumulative exposure measure. The 8-year value may better reflect nonadherence to lipid lowering treatments or worsening social conditions in older age, which is highly associated with stroke, though reasons for this pattern of association remain unclear and require further research on mechanisms for stroke development in African American adults.

We also observed associations between cumulative FPG exposure and HF and stroke. While hyperglycemia is a well-established risk factor for microvascular complications and diabetes, its cumulative impact on macrovascular outcomes has been less studied, particularly in African American populations. One study examining the cumulative exposure to hemoglobin A1c levels >7% over a mean 5 years found a modestly increased association with CVD events compared to using a single value, but this study did not account for exposure to pre-diabetic A1c levels or include African American men and women.^24^ Hemoglobin A1c may also underestimate hyperglycemia in African American people due to the prevalence of sickle cell trait, potentially making FPG a more accurate longitudinal measure in this group.^25^ From a biological perspective, cumulative FPG levels reflect the direct impacts of hyperglycemia on vasculature, after adjustment for non-glycemic effects of diabetes (e.g, dyslipidemia, obesity). In our study, the estimated effect of cumulative FPG may reflect a combination of those pathways due to residual confounding or interaction, even after adjustment for diabetes status and other risk factors; despite this, the observed associations of both the cumulative and proximal values of FPG with HF and stroke point to the importance of managing blood glucose levels and warrant further investigation into the mechanisms of hyperglycemia on CVD risk in African American populations.

### Strengths and Limitations

We considered multiple cumulative exposure calculations in this study, including areas above clinically recognized harmful thresholds, and found that higher total exposure measures resulted in the strongest associations with CVD events across risk factors. This suggests that exposure to subclinical levels of risk factors confers additional CVD risk,^3,26^ and that measures that do not account for subclinical exposures may underestimate risk. While this total exposure model may be appropriate for LDL-C, it may not generalize in all cases for SBP or FPG, where disordered states exist at low exposure levels (i.e., hypotension and hypoglycemia). In our study, these levels were rare, and we were unable to account for a J- or U-shaped association between risk factor levels and outcomes.

The current study is also subject to other limitations. First, the use of only three examinations limits the precision of cumulative exposures and may underestimate variability in the exposures. Second, follow-up after the exposure period limited the number of CHD and stroke events, thus reducing statistical power to detect associations. Third, we note that the 8-year window does not capture lifetime exposure, which is likely even more relevant to long-term CVD risk. However, despite these limitations, our results support the use of cumulative exposure measurements over single values. Fourth, there is a possibility of chance findings as we performed multiple statistical comparisons without multiplicity adjustment; therefore, all reported 95% confidence intervals may be too narrow and should be interpreted jointly with point estimates. Fifth, there is potential for misclassification of the 8-year value due to intercurrent events and interpolation of the linear trajectory from Exam 2 to Exam 3; however, this is likely minimal as the proportion of events occurring prior to Exam 3 is low (3% of CHD, 2% of HF, and 2% of stroke). Finally, exclusion of participants without Exam 3 measurements (40% of the baseline cohort) may have attenuated observed associations, if missing values are associated with both higher cumulative exposures and higher outcome rates.

### Perspectives

Higher cumulative exposure to traditional CVD risk factors was associated with CVD outcomes in African American adults; higher cumulative SBP and LDL-C were associated with CHD and higher cumulative SBP and FPG were associated with HF. These findings emphasize the importance of early detection, sustained control, and regular monitoring of SBP, LDL-C, and FPG to reduce lifetime CVD risk. Interventions aimed at lowering and managing exposure to risk factors over the life course rather than solely treating risk factors once they are markedly elevated may represent effective strategies for reducing the burden of CVD in African American adults.

## Disclosures

The Jackson Heart Study is supported by Contracts HHSN268201800010I, HHSN268201800011I, HHSN2628201800012I, HHSN2628201800013I, HHSN2628201800014I, HHSN2628201800015I from the National Heart, Lung, and Blood Institute (NHLBI) with additional support from the National Institute on Minority Health and Health Disparities (NIMHD). This manuscript was supported by grant R01AG071019 from the National Institute on Aging (NIA).

The views expressed in this manuscript are those of the authors and do not necessarily represent the views of the NHBLI, the NIMHD, the National Institutes of Health, or the US Department of Health and Human Services.

This manuscript has been reviewed by JHS for scientific content.

## Data Availability

Jackson Heart Study data may be requested from the Biologic Specimen and Data Repository Information Coordinating Center (BioLINCC) repository and the database of Genotypes and Phenotypes (dbGaP). Additionally, investigators with a manuscript proposal or ancillary study proposal that has been approved by study committees may request data directly from the Jackson Heart Study Coordinating Center. BioLINCC: https://biolincc.nhlbi.nih.gov/studies/jhs/ dbGaP: https://www.ncbi.nlm.nih.gov/gap/ Jackson Heart Study Coordinating Center: https://www.jacksonheartstudy.org/Research/Study-Data/Data-Access

https://www.ncbi.nlm.nih.gov/gap/

https://biolincc.nhlbi.nih.gov/studies/jhs/

